# The effect of COVID-19 on the economy: evidence from an early adopter of localized lockdowns

**DOI:** 10.1101/2020.09.21.20198887

**Authors:** Kenzo Asahi, Eduardo A. Undurraga, Rodrigo Valdés, Rodrigo Wagner

## Abstract

**Background:** Governments worldwide have implemented large-scale non-pharmaceutical interventions, such as social distancing or school closures, to prevent and control the growth of the COVID-19 pandemic. These strategies, implemented with varying stringency, have imposed substantial social and economic costs to society. As some countries begin to reopen and ease mobility restrictions, lockdowns in smaller geographic areas are increasingly being considered as an attractive policy intervention to mitigate societal costs while controlling epidemic growth. However, there is a lack of empirical evidence to support these decisions.

**Methods:** Drawing from a rich dataset of localized lockdowns in Chile, we used econometric methods to measure the reduction in local economic activity from lockdowns when applied to smaller or larger geographical areas. We measured economic activity by tax collection at the municipality-level.

**Findings:** Results show lockdowns were associated with a 10-15% drop in local economic activity, a two-fold reduction compared to municipalities not under lockdown. A three-to-four-month lockdown had a similar effect on economic activity than the year of the 2009 great recession. We found that costs are proportional to the population under lockdown, without differences when lockdowns were measured at the municipality or city-wide levels.

**Conclusions:** Our findings suggest that localized lockdowns have a large effect on local economic activity, but these effects are proportional to the population under lockdown. Our results suggest that epidemiological criteria should guide decisions about the optimal size of lockdown areas; the proportional effects of lockdowns on the economy seem to be unchanged by scale.

**JEL codes:** I10, I15, I18, H2

## Introduction

Non-pharmaceutical interventions are still the main strategies to control viral transmission in the COVID-19 pandemic [1-3]. These interventions range from individual-level recommendations, such as the use of facemasks or frequent hand-washing, to large-scale regulations and policies, such as large-scale quarantines or lockdowns and non-essential business closures [4,5]. Several countries have achieved some control over the COVID-19 based on a combination of non-pharmaceutical interventions and high levels of testing and isolation of infected people [6-12]. However, there is a substantial risk of a resurgence of the epidemic [13-15]. Understanding the impacts of these interventions is critical because they will most likely continue until an effective vaccine becomes available for a substantial proportion of the population [16]. There is still limited empirical evidence of the effects of interventions to prevent viral transmission [1,17]; most impact has been estimated using mathematical models [16,18,19]. The COVID-19 pandemic has already imposed an enormous global burden, with about 30 million cases and one million deaths reported so far [20], and substantial social and economic costs from epidemic control measures [21-27].

As countries begin to reopen and ease mobility restrictions, localized lockdowns are increasingly considered a critical element of a non-pharmaceutical toolkit to control COVID-19 resurgence [6,7,19,28,29]. In contrast to nation-wide lockdowns, localized lockdowns are implemented over a limited geographical area, ranging from a neighborhood to a city, including suburbs, districts, or towns. Localized lockdowns are appealing to policy-makers because, in principle, they would allow countries to reopen and reclose specific jurisdictions based on local virus transmission dynamics. Large-scale lockdowns are unsustainable because of the high costs they impose on the population [12]. Thus, compared to large-scale interventions, localized lockdowns may control transmission hotspots while mitigating some social and economic costs. Policy-makers need to make decisions about COVID-19 control strategies, considering their epidemiological, social, and economic effects.

Epidemiological evidence is one amongst several criteria for decision-making regarding non-pharmaceutical interventions. For example, a policy-maker would want to understand if costs of foregone economic activity are proportional to the population under lockdown, or whether costs are mitigated or amplified when lockdowns are implemented at different administrative levels (e.g., municipality, city, state, country). On the one hand, demand spillovers would suggest that people in a municipality could buy in stores of the neighboring municipality and, through substitution, limit the economic fallout in the city as a whole. On the other hand, the fall in economic activity could be more than proportional if a lockdown affects supply chains, such as when workers cannot work in a neighboring municipality because of mobility restrictions in their municipality of residence. The answer to this question is mostly empirical, as there are good arguments to both sides. However, there is limited and non-conclusive evidence on the economic costs of non-pharmaceutical interventions. Researchers in the United States have examined how non-pharmaceutical interventions have impacted unemployment insurance, employment, or store client traffic. Some research suggests that lockdowns explain a small share of the total economic activity decline [22,30-32]. Others [24,33,34] have found that lockdowns play a relevant role in explaining the drop in economic activity. We test these effects in a setting were localized lockdowns were implemented intermittently at different administrative levels, thus allowing us to identify the impact of localized lockdowns on economic activity.

The World Health Organization declared South America as the new epicenter of COVID-19 on May 22, 2020 [35]. Despite implementing several epidemic control strategies early in the pandemic, including travel restrictions, school closures, and mandatory lockdowns [36], the pandemic has imposed a massive toll in the region. As of September 13, South America has reported more than 240 thousand deaths; Colombia, Brazil, Peru, and Chile are among the ten countries with most reported COVID-19 infections globally and the spread is far from controlled [20,37,38]. While mostly failing to stop viral spread [37,39], Latin America is now facing the social and economic costs of large-scale non-pharmaceutical interventions. Since the beginning of the epidemic, Chile has implemented localized lockdowns at the municipality level, the smallest administrative subdivision in the country, at various points in time (Figure 1). The government roughly defined the criteria for implementing localized lockdowns as a function of disease burden, growth in transmission, and healthcare capacity, but did not define specific thresholds [40]. Epidemiological evidence suggests localized lockdowns reduce epidemic growth[41], but are heavily affected by spillovers from neighboring communities [17] and income [42,43]. Localized lockdowns have helped contain the transmission of the virus in isolated areas. Still, they cannot control the epidemic in highly interdependent areas, such as municipalities within a metropolitan area.

**Figure 1.**
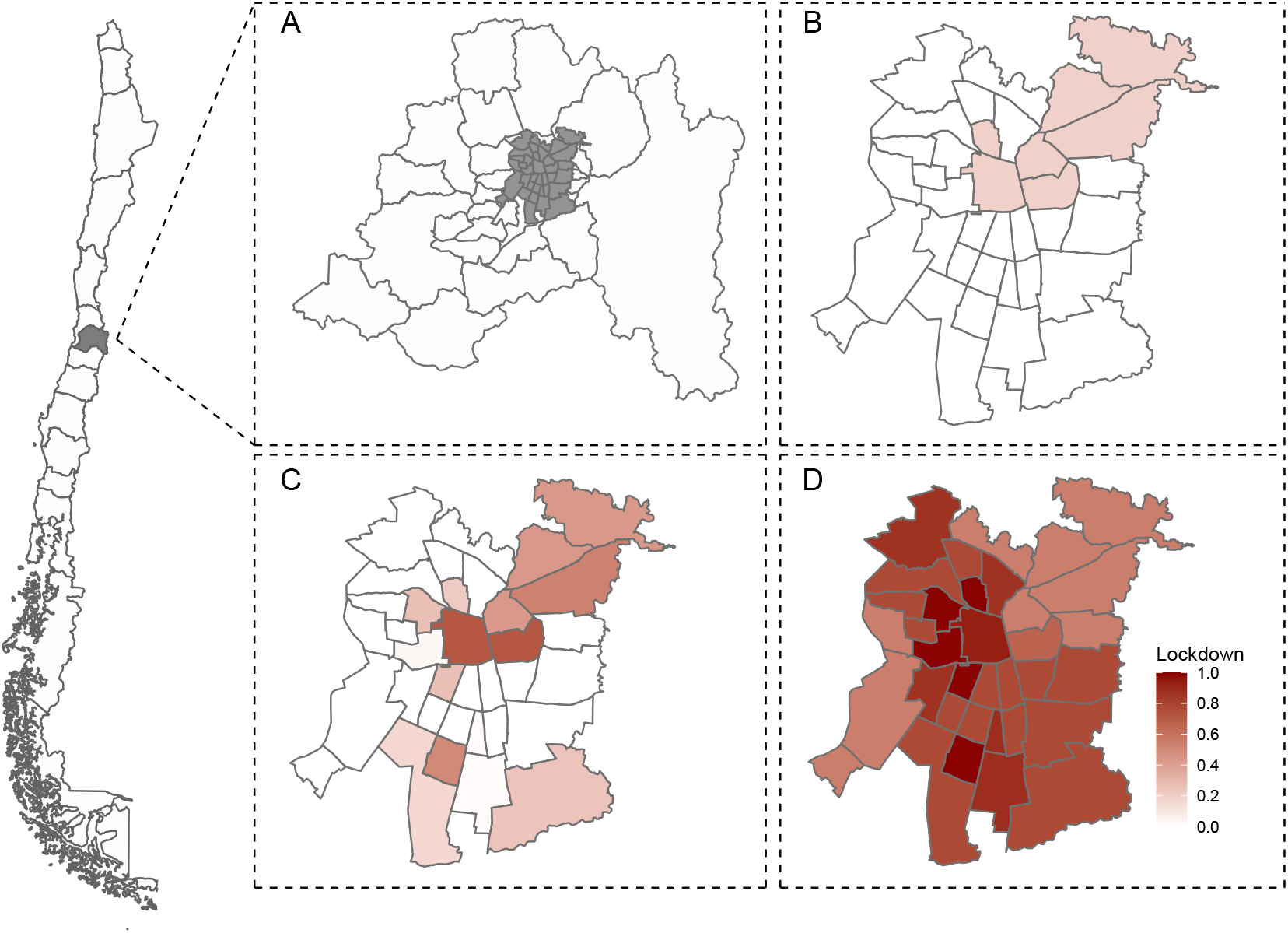
Illustration of localized lockdowns at the municipality leve, Greater Santiago, Chile, March-May 2020. To control COVID-19 growth, the Ministry of Health implemented localized lockdowns at the municipality level, the smallest administrative subdivision in the country. The figure illustrates these lockdowns implemented in Greater Santiago (A in gray), and different points in time: March 30 (B), April 30 (C), and May 30 (D). Localized lockdowns were implemented at the municipality-level across the country.

Drawing on a rich integrated dataset, including value-added tax (VAT) revenues, population data, and daily incidence of lab-confirmed COVID-19 cases, we use econometric methods to empirically estimate the economic costs of these localized lockdowns in Chile. We hope these results will help inform policy implementation decisions in the context of the COVID-19 pandemic.

## Methods

### Data

Value-added tax (VAT) applies to all goods with a flat rate of 19% in Chile. VAT is collected and paid monthly to the Chilean tax authority (Servicio de Impuestos Internos). Our data includes VAT at the municipality level, by all firms registered in the Chilean tax authority, for 2018-2020. VAT collection has a tight one-to-one relationship with GDP; it is, therefore, a good proxy for economic activity. Both variables cointegrate in time series and panel analysis; error correction models suggest that half-life deviations vanish in less than a year [44].

We used Chile’s 2017 National Census [45] to estimate the population in each municipality, and epidemiological surveillance records for COVID-19 from Chile’s Ministry of Health [40,46]. We obtained mobility data from the Data Science Institute at Universidad del Desarrollo [47]. Data on COVID-19, mobility, and population are publicly available on institutional websites [45-47]. The data on VAT used for this study is available from the corresponding author upon reasonable request and with permission of the tax authority.

### Analysis

We used the collection of the VAT as our dependent variable. Our lockdown variable corresponds to the proportion of days that a municipality *i* is in quarantine in a given month *t*:

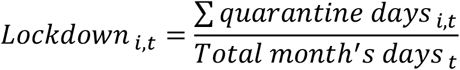

We limited our analysis of the 170 municipalities with above-median total VAT in 2018, excluding mostly small and rural municipalities. This preferred sample of municipalities includes 97% of Chile’s 2018 VAT and 89% of the population (Figure 2). Our sample also excluded the three municipalities that concentrate large-company headquarters (Santiago, Las Condes, and Providencia), such as banks and mining companies, because VAT data in these municipalities do not reflect local economic activity.

**Figure 2.**
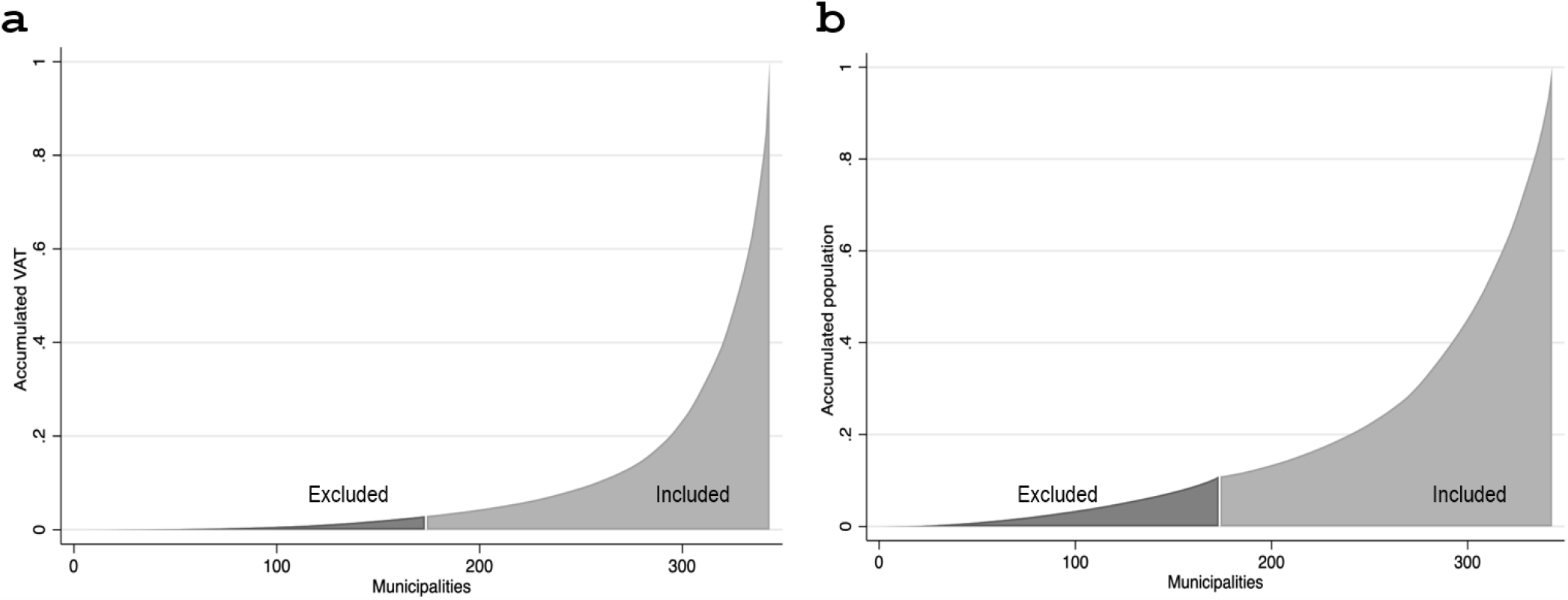
VAT and population cumulative distribution across all municipalities. Panel (a) shows the proportion of total 2018 VAT considered in our baseline sample. We sorted the 343 municipalities in our dataset in ascending order by 2018 VAT, and we calculated the accumulated tax from the one with the lowest to the one with the highest level of VAT. Municipalities not considered in our baseline sample account for 2.9% of the total 2018 VAT (darker area), while the remaining 97.1% (lighter area) is in our preferred sample. Panel (b) shows the proportion of the total population according to the 2017 Census within our preferred sample. In this case, we sorted the municipalities in ascending order. We then calculated the accumulated percentage of the total population that is not considered in our sample, which is 10.9% (darker area). Hence, the remaining 89.1% (lighter area) is in our sample.

Our main empirical specification is a two-way fixed-effects model:

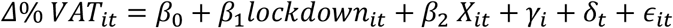

where *Δ%VAT*_*it*_ corresponds to the percent variation of total VAT collected in municipality *i* at month *t* in 2020 relative to the same month in 2019. *lockdown*_*it*_ is our variable of interest and represents the proportion of days in a month that a municipality was under lockdown. *γ*_*i*_ and *δ*_*t*_ correspond to municipality and time fixed-effects, respectively. A distinctive feature of our setting is that *lockdown*_*it*_ effectively changes by municipality and month, providing a variation that allows for a plausible estimate of effects (Figure 1). We controlled for threat or risk perception [48] and social distance by adding COVID-19 cases or deaths in the municipality *i* at time *t* (variable X_it_) as a covariate. For instance, people may not open their businesses or spend in the local economy because they fear COVID-19 contagion, independent of whether their municipality is under lockdown or not.

Similar to virus transmission spillovers [17], the economic effects of localized lockdowns within a city or in a conurbation may differ from more relatively isolated municipalities with no neighboring urban areas (“standalone” municipalities). To examine whether the impact of lockdowns on economic activity is heterogeneous depending on whether municipalities belong to a conurbation or are a standalone municipality, we used the following regression specification:

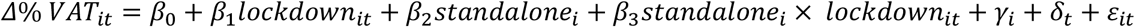

where *standalone*_*i*_ takes a value of one for standalone municipalities and zero otherwise.

The economic effects of localized lockdowns may also differ depending on the area under lockdown—for example, at the municipality or conurbation level. To examine this question, we also ran our analysis comparing all municipalities within a conurbation with standalone municipalities. We weighted the number of days in quarantine in month *t* of each municipality *i* belonging to the conurbation *c* according to the total 2018 VAT:

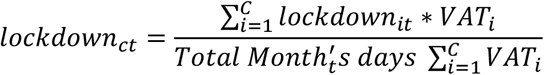

We estimated deaths and COVID-19 cases as a weighted average of deaths in municipalities within the conurbation, using the municipality’s population as the weight. Hence, the equation describing per capita COVID deaths in each conurbation is as follows:

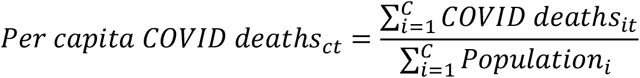

Last, we investigate how mobility at the municipality-level affects economic activity. We used a mobility index based on cellphone data.

## Results

### Descriptive statistics

Table 1 shows the main descriptive statistics of our sample. Figure 3 shows the longitudinal effects of lockdown. Municipalities without a localized lockdown saw a 15% drop in VAT collection in April-May. By contrast, municipalities in lockdown suffered a more massive decline of 25-30% in VAT collection. Figure 4 shows a cross-section, considering month and municipality fixed-effects. The figure shows a clear relationship between the extent of lockdowns and the decline in VAT.

**Table 1.**
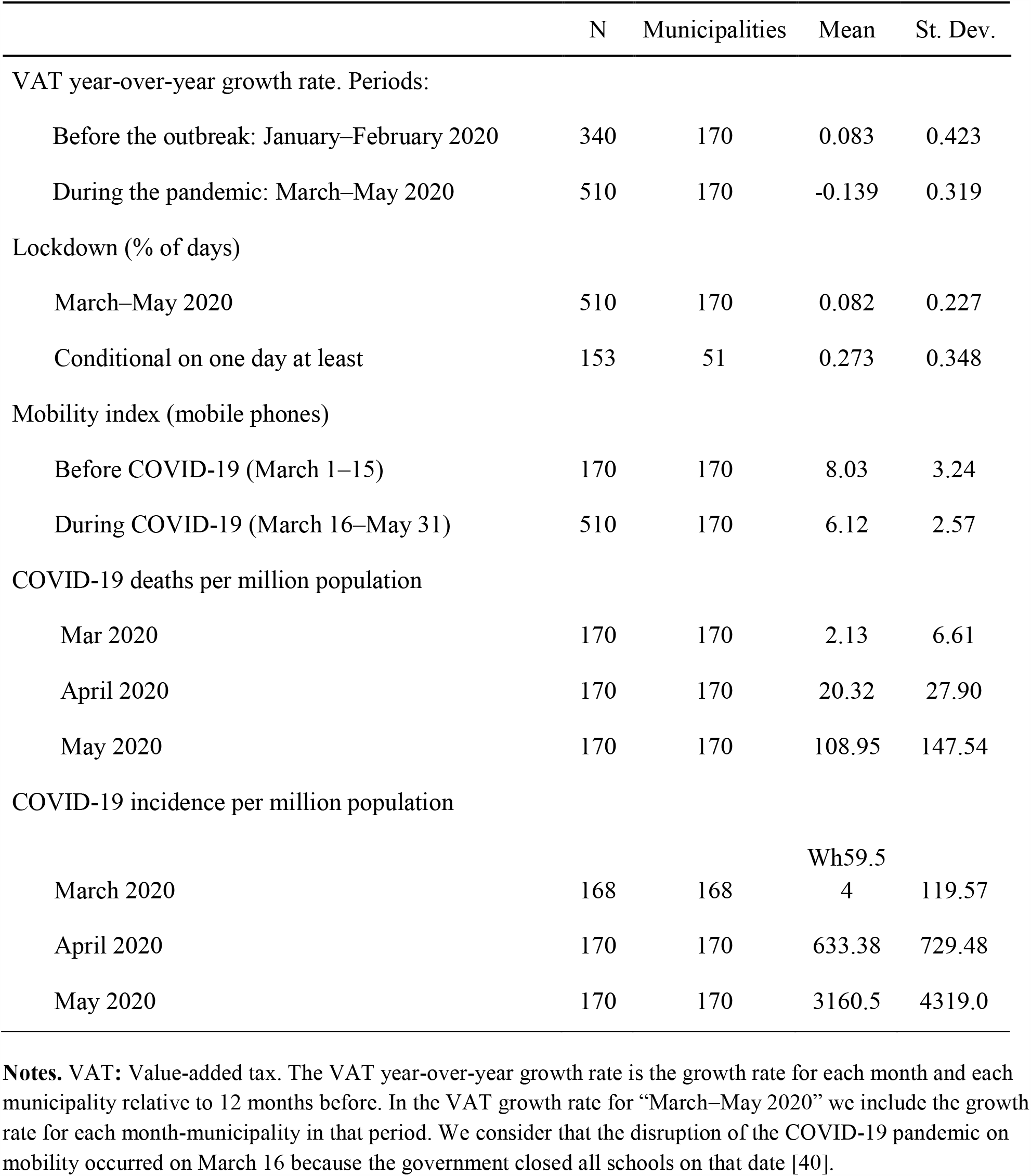
Descriptive statistics related to localized lockdowns in Chilean municipalities, March-May 2020

**Figure 3.**
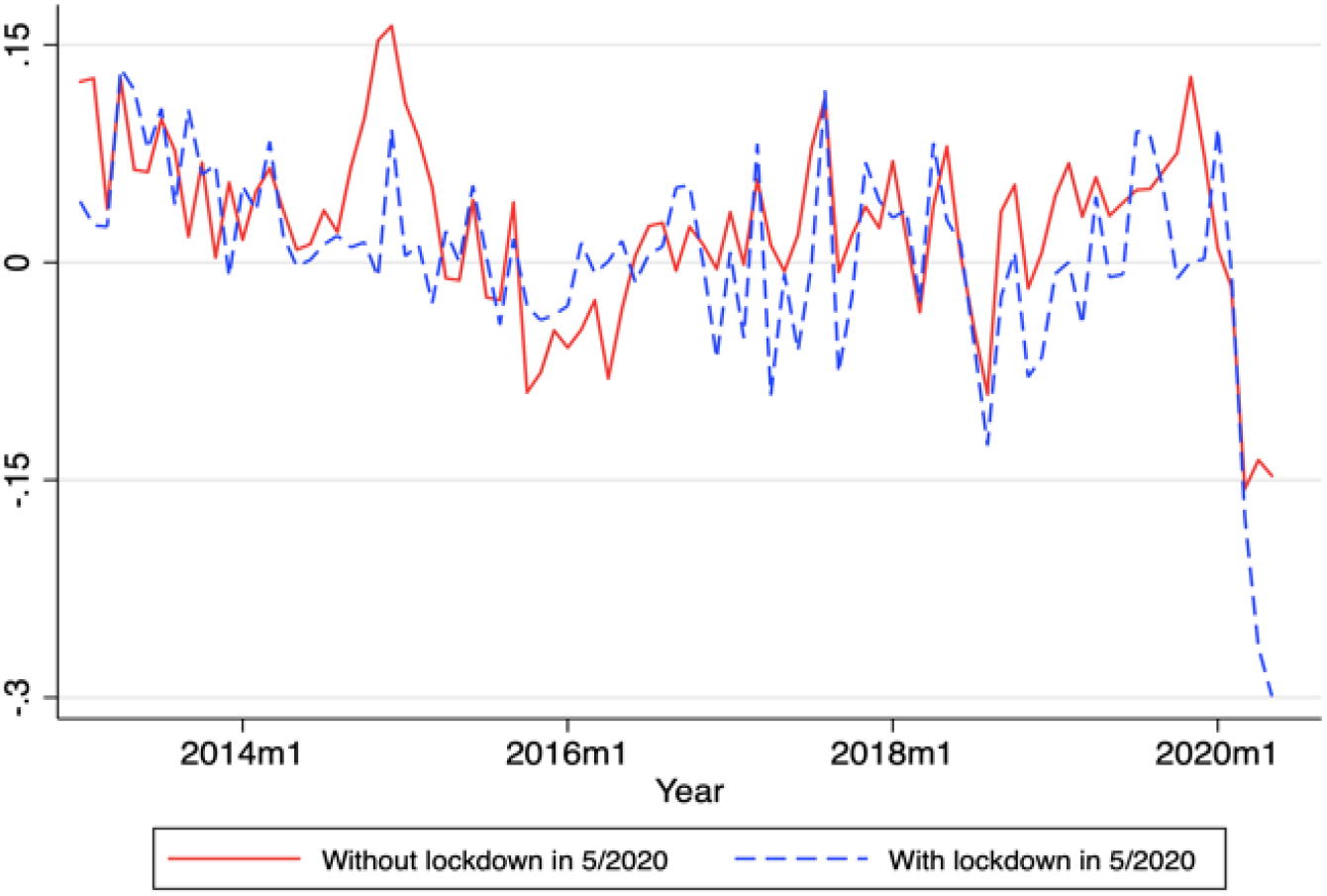
Median of the real value-added-tax (VAT) year-on-year growth rates. The graphs shows the median of VAT growth rates for municipalities that were under lockdown in May 2020 (blue) and municipalities that were not under lockdown (red). The median of the value-added-tax (VAT) growth rate in May 2020 for municipalities with and without lockdown is 2.67 and 5.37 standard deviations lower than the mean of such medians in the 2006-2019 period. The sample of municipalities includes municipalities over the 50th percentile of the total 2018 VAT.

**Figure 4.**
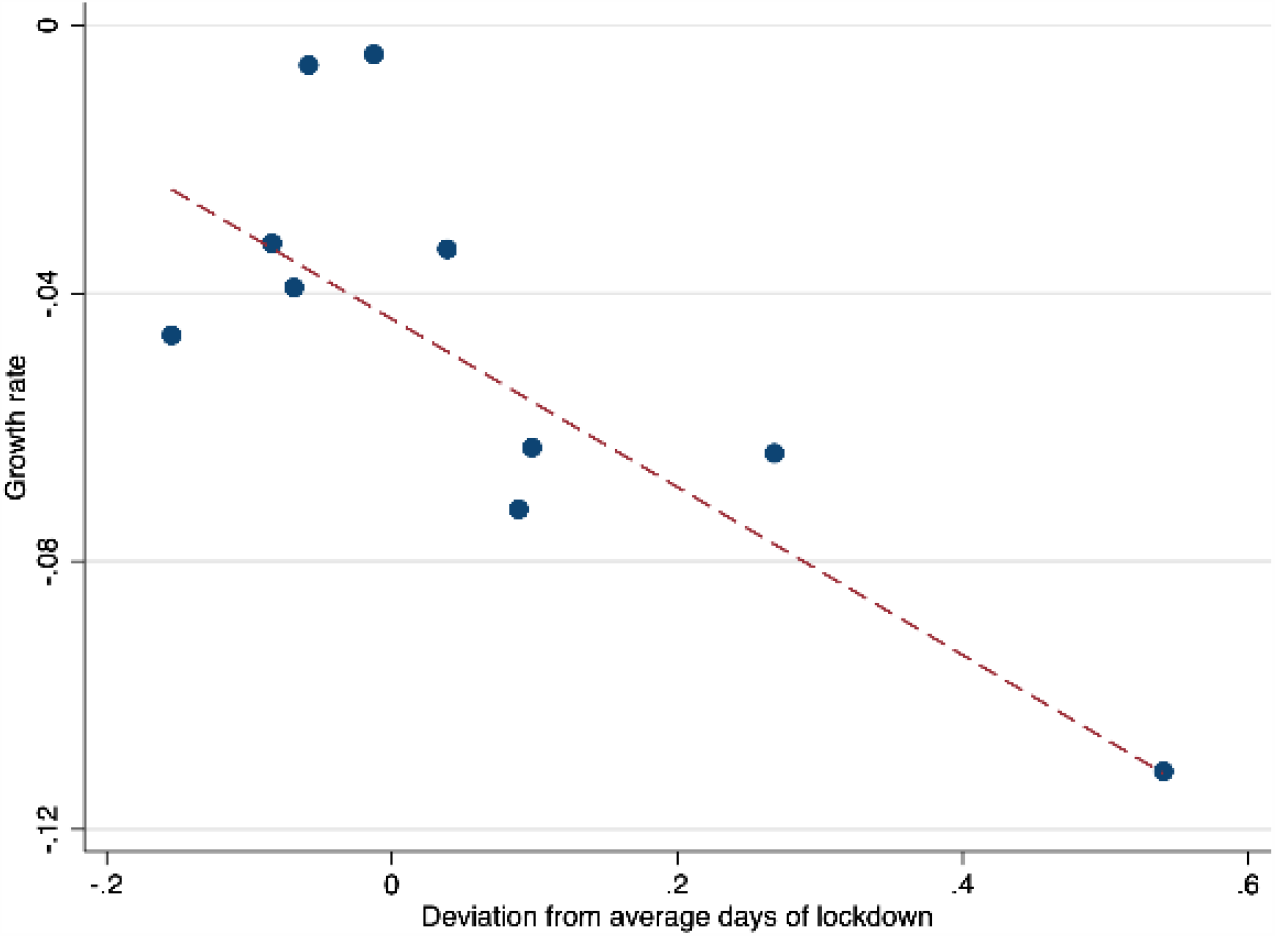
Effect of lockdown on value-added-tax (VAT) collection for January 2020 through May 2020, controlling for month and municipality fixed effects. The results show the association between lockdown on VAT collection for January 2020 through May 2020, controlling for month and municipality fixed effects. We group the municipalities of our baseline sample into equal-sized bins according to days of lockdown between January 2020 and May 2020. Each dot represents the mean of VAT collection growth rate (y-axis) and the mean of deviation from lockdown as a percentage of a month (x-axis), within each bin. Each bin has 17 municipalities. The red dashed line represents the population regression line.

### Multivariate analysis

#### Municipality level

Table 2 presents our baseline results for the effect of lockdowns on economic activity. Table 2, column

**Table 2.** Regressions results for the effect of one month localized lockdown on total VAT collection, estimated with two-way fixed effects at the municipality level, January-May 2020

| Dependent variable:<br>VAT growth | Baseline | Excluding rural units | Excluding Greater Santiago | Conurbations | Conurbations excluding Greater Santiago | Conurbations and standalone municipalities | As in (1) controlling for deaths | As in (1) controlling for cases |
| --- | --- | --- | --- | --- | --- | --- | --- | --- |
|  | (1) | (2) | (3) | (4) | (5) | (6) | (7) | (8) |
| Lockdown | -0.125***<br>(0.048) | -0.132***<br>(0.049) | -0.162*<br>(0.096) | -0.161**<br>(0.064) | -0.153<br>(0.130) | -0.230***<br>(0.059) | -0.125*<br>(0.071) | -0.135***<br>(0.052) |
| Standalone × lockdown |  |  |  |  |  | -0.059<br>(0.104) |  |  |
| Standalone |  |  |  |  |  | -0.005<br>(0.038) |  |  |
| Death per 100,000 population |  |  |  |  |  |  | 0.00004<br>(0.002) |  |
| Log Incidence per 100,000 population |  |  |  |  |  |  |  | 0.002<br>(0.005) |
| Observations | 850 | 785 | 570 | 360 | 195 | 455 | 850 | 850 |
| Adjusted R<sup>2</sup> | 0.352 | 0.369 | 0.356 | 0.352 | 0.278 | 0.171 | 0.351 | 0.351 |
| Time effect | YES | YES | YES | YES | YES | YES | YES | YES |
| Municipalities | 170 | 157 | 114 | 72 | 39 | 91 | 170 | 170 |

**Notes.** All specifications have both geography and time fixed effects. VAT: Value-added tax. Robust standard errors in parenthesis. Column (1) shows regression results for the baseline sample (i.e., municipalities over 50th percentile of total 2018 VAT). Column (2) excludes units with over fifty percent of rural population. Column (3) is the same as (2) but excluding municipalities in Greater Santiago, the capital. Columns (4) includes only municipalities that are part of a large conurbation. Column (5) is the same as column (4), excluding municipalities in Greater Santiago. Column (6) includes municipalities that are part of large conurbations and standalone municipalities (Angol, Antofagasta, Arica, Aysén, Calama, Castro, Chañaral, Colina, Copiapó, Curicó, Osorno, Ovalle, Puerto Montt, Puerto Natales, Punta Arenas, Valdivia, and Vallenar). Columns (7) and (8) consider the baseline sample and controls for contemporaneous COVID-19 deaths and incidence per 100,000 population, respectively, at the municipality level.

(1) shows that one month of lockdown decreases monthly VAT around 12.5% (β:-0.125; 95% CI:-0.220,-0.031; p<0.01). The coefficient or the effect of lockdowns has about the same magnitude when restricting the sample to municipalities with at least 50% of the urban population (Table 2, column 2; β: −0.132; 95% CI:-0.228,-0.035; p<0.01). Table 2, column (3) shows the results for municipalities with less than 50% of the rural population and excluding observations from Greater Santiago. To assess the robustness of our estimates, we excluded municipalities in Greater Santiago, the conurbation in Chile with the highest proportion of municipalities in lockdown between March and May 2020. We found that VAT decreases 16 percentage points for each month of lockdown, but the coefficient is only significant at the 90% level (β=-0.162; 95% CI:-0.350,0.0268; p<0.10).

.We then limited our sample to urban municipalities (n=72) that are part of a conurbation (Table 2, column 4). One month of lockdown results in a monthly VAT decrease of 16 percentage points (β=-0.161; 95% CI:-0.287,-0.034; p=0,013). We found similar results when excluding Greater Santiago (Table 2, column 5; β=-0.153; 95% CI:-0.410,0.103; p=0.240).

We added an interaction term to examine whether lockdowns had a different effect on VAT in municipalities that are part of a conurbation or in standalone municipalities. The results in Table 2, column (6) show a 23% decline in monthly VAT collection due to a one-month lockdown (β=-0.230; 95% CI:-0.345,-0.115; p<0.01). However, we did not find evidence of a differential effect for standalone municipalities relative to municipalities in conurbations.

Last, we examined whether perceived threat or risk from COVID-19 deaths or cases could be an omitted variable bias in the effect of local lockdowns on economic activity. Table 2, Column (7) includes the municipality’s one-month lagged per-capita COVID deaths per 100,000 population as control. The effect of lockdown is roughly the same as in column 1 (β=-0.125; 95% CI:-0.265,0.013; p<0.10). Controlling for COVID-19 monthly incidence per 100,000 population Table 2, Column (8) shows that one month of lockdown results in a thirteen percent decrease in VAT collection (β=-0.135; 95% CI:-0.237,-0.033; p=0.01). Results are robust to using one-month lagged COVID-19 deaths and cases.

Overall, Table 2 suggests that one month of lockdown would reduce economic activity by 10-15%, robust to several model specifications. Notably, the effect size is not affected when controlling for COVID-19 deaths or case incidence, suggesting that the lockdown effect in this sample goes over and above the impact of perceived threat or risk of contagion.

#### Conurbations and standalone municipalities

Next, we examined the effects of lockdowns on VAT when analyzed for conurbations or standalone municipalities (Table 3). The objective was to test whether the effects of lockdowns were different when there were no spillovers from closely interdependent neighboring areas. For the analysis, we collapsed municipalities into conurbations. Our sample now had eighteen conurbations and seventeen standalone municipalities in our baseline sample.

**Table 3.** Regressions results for the effect of one month localized lockdown on total VAT collection, estimated with two-way fixed effects for conurbations and standalone municipalities, January-May 2020

| Dependent variable:<br>VAT growth | All<br>Conurbations<br>and standalone<br>municipalities | Excluding<br>Greater<br>Santiago | As in (1)<br>interacting<br>lockdown &<br>standalone | As in (1)<br>with per<br>capita<br>deaths | As in (1)<br>with log per<br>capita<br>incidence |
| --- | --- | --- | --- | --- | --- |
|  | (1) | (2) | (3) | (4) | (5) |
| Lockdown | -0.184**<br>(0.089) | -0.188*<br>(0.020) | -0.243***<br>(0.064) | -0.126<br>(0.106) | -0.157<br>(0.116) |
| Standalone |  |  | -0.00294<br>(0.044) |  |  |
| Standalone × lockdown |  |  | -0.042<br>(0.105) |  |  |
| Lagged per capita deaths<br>per 100,000 population |  |  |  | -0.007<br>(0.006) |  |
| Log incidence<br>per 100,000 population |  |  |  |  | -0.014<br>(0.027) |
| Observations | 175 | 170 | 175 | 175 | 175 |
| Adjusted R<sup>2</sup> | 0.325 | 0.322 | 0.101 | 0.326 | 0.323 |
| Units | 35 | 35 | 35 | 35 | 35 |
| Conurbations | 18 | 18 | 18 | 18 | 18 |
| Standalone municipalities | 17 | 17 | 17 | 17 | 17 |

**Notes.** All specifications have both geography (conurbation, standalone municipalities) and time fixed-effects. VAT: Value-added tax. Robust standard errors in parenthesis. Localized lockdowns at the conurbation level are aggregated and weighted by total 2018 VAT. COVID-19 deaths and cases are calculated at the conurbation level.

Table 3, column (1), shows a statistically significant decline in monthly VAT collection of around 18% (β=-0.184; 95% CI:-0.360,-0.009; p<0,050). Because Greater Santiago had the largest number of municipalities with lockdown, we dropped Greater Santiago from the sample to test our results (Table 3, column 2). The magnitude of the effect remained but was significant only at the 90% level (β=-0.188; 95% CI:-0.382, 0.051; p=0,056). In Table 3, column 3, we examined whether there was a differential effect for standalone municipalities. The results show that one month of lockdown results in a significant decrease of 24% of VAT collection (β=-0.243; 95% CI:-0.370,-0.117; p<0,001); we did not find evidence for a differential effect in standalone municipalities. However, the coefficient in Table 3, column 3 was not statistically different from the coefficient in Table 3, columns (1) and (2).

Last, we examined whether the lockdown effect was different from the perceived threat or risk from COVID-19. In Table 3, columns (4) and (5) show lockdowns were no longer statistically significant at conventional levels (p=0.240 and p=0.175, respectively). However, the coefficient’s sign was still negative and about the same magnitude as the coefficient in Table 3, columns (7) and (8). The joint significance test for the proportion of the month under lockdown and lagged per capita COVID-19 deaths and incidence was significant (F=3.84, p<0.05; F=2.81, p=0.064, respectively). Thus, working with data at the conurbation-level instead of the municipality-level makes it harder to disentangle the effect of lockdowns. This is partly explained by insufficient statistical power and, from limited variation in the lockdown variable. The last columns of Table 3 reinforce the advantage of our baseline setting at the municipal level, with more sizable variation in the lockdown (key) variable.

Table A1 in the Supplemental material also shows that our baseline results are robust to controlling for a measure of cellphone-based mobility. However, we also argue that it might be misleading to control for mobility since it is one of the main mediating channels by which lockdown affects economic activity (see Supplemental material for further discussion).

## Discussion

Our results suggest that a full-month lockdown explains a drop in activity of the order of 10-15 percentage points, almost twice the reduction for non-locked down areas. While the expected sign of the effect of lockdowns on economic activity might be obvious, its magnitude is not.

These estimates are large. Our estimates suggest that a three-to-four-months lockdown would reduce economic activity by approximately the same amount that the recession affected the Chilean economy in the (whole) year 2009. During the 2009 Great Recession, GDP declined by 1.1% instead of growing by 3.7% [49]. These three to four months only consider the additional effect of lockdowns. If one considered the whole drop in economic activity, the magnitude would be twice as much (in two months under lockdown in 2020 the drop in GDP is comparable to the annual drop in 2009).

Another way of thinking quantitatively about the magnitude and implications of our baseline estimate in terms of employment. Assuming a standard short-run labor-to-economic activity elasticity of around 0.3-0.5, as suggested by an OECD study [34], a one-month lockdown would imply a drop of about 6% in monthly employment. We estimate this illustrative 6% fall in monthly employment, by multiplying the coefficient of −0.15 in Table 2 by an average short-run labor-to-economic activity elasticity of 0.4.

It is also useful to contrast our results with the polar case of South Korea, without lockdowns. Aum et al. [24] found that a one-per-thousand increase in the infection rate was associated with an employment loss of 2-3%. Extrapolating this result to the United States and the United Kingdom, which had large-scale lockdowns, Aum et al. [24] argue that only half of the 5-6% drop in job losses in these Western economies might be attributable to lockdowns. The rest would be from social panic, some other large-scale non-pharmaceutical intervention, such as school closures, or demand effects. This similar effect of areas with and without lockdown seems consistent with our findings. Importantly, we obtained our results from a direct test in the same sample, instead of extrapolating across countries. The relatively large effect of lockdowns has not yet been found empirically in the United States. For instance, Bartik et al. [30] found that the relative impact of lockdowns was smaller, explaining 1/6 of the total fall during the COVID-19 pandemic. Our results show that lockdowns explain half of the effect, both in the raw time series (Figure 3) and in the main regressions (Table 2). Thus, we offer a qualification to Brzezinksi et al. [50], who found that not imposing lockdowns barely improves economic performance, while drastically increasing medical costs. This baseline drop probably includes threat or risk perception and includes other economic channels, like lower spending [25].

Epidemiological evidence suggests localized lockdowns reduce epidemic growth [41], but their effectiveness is affected by spillovers from neighboring areas where there is economic interdependence, such as in a city [17]. From an epidemiological standpoint, governments should implement localized lockdowns at the city-level, where “buffer” zones exist to minimize transmission networks [28]. We examined localized lockdowns at different scales to understand their relative economic costs, understanding that this is only one portion of the relevant cost calculation. Our findings suggest no disproportionate economic gains from unlocking a part of the city. Our estimated effects of lockdowns on the economy are unchanged by scale. The plausible channels that mitigate or amplify the economic impact in the case of a widespread vis-à-vis a local lockdown do not seem critical, at least in our study setting, enforcing the convenience of implementing localized lockdowns at the city or commuting area levels.

Economic problems could also feedback into health through several channels. For instance, a drop in economic activity of 10-15 percentage points is relevant because lockdowns can affect government budgets, even in the long term. For example, Frenier et al. [51] argue that several states in the USA will probably face severe budget deficits from reductions in tax revenue from the pandemic. Further, an economic downturn may prompt many deaths of despair and mental illness from unemployment and isolation [52].

Our estimates have limitations. First, we used a tax payment as a proxy for economic activity. Nonetheless, we also have VAT and survey-based employment at the regional level in Chile. We found a statistically significant elasticity of 0.3 between the drop in VAT and the decline in total employment (including self-employed), consistent with short-run output-employment elasticities in the literature (34). Another limitation is that informal economic activity is, by its very nature, not directly captured in our measures of VAT. This is a more general limitation globally. However, compared to Latin America, Chile has relatively low informality [53].

Our study may also have other confounders. For instance, the government gave some leeway on when to pay taxes, and we could only examine monthly-level observations. Nevertheless, there are no apparent reasons why these confounders may interact with lockdowns. These confounders may have also introduced measurement error in our tax measures. This would have increased our standard errors, making it more difficult to get statistical significance. Nevertheless, we did get relevant and robust estimates across various specifications, which mitigates the concerns related to measurement error.

## Conclusions

We used a rich dataset of localized lockdowns in Chile to measure their effect on economic activity. We find sizeable impacts of lockdowns, doubling the drop in economic activity compared to non-treated municipalities, and robust to several model specifications and controls. As many countries are beginning to reopen and ease mobility restrictions, localized lockdowns can be a critical tool to control COVID-19 resurgence while minimizing economic impact. We found no evidence that localized lockdowns generate a proportionally larger or smaller effect in the economy when applied to areas of different sizes.

Critically, our results suggest that epidemiological criteria should guide decisions about the optimal size of lockdown areas; the proportional effects of lockdowns on the economy seem to be unchanged by scale.

## Data Availability

Data on COVID-19, mobility, and population are publicly available on institutional websites. The data on VAT used for this study is available from the corresponding author upon reasonable request and with permission of the tax authority.

http://www.minciencia.gob.cl/covid19

https://www.censo2017.cl/

https://www.gob.cl/coronavirus/cifrasoficiales/

## Acknowledgments

We thank Sebastian Piña (Universidad de Chile) for research assistance, Paula Aguirre (Pontificia Universidad Católica de Chile) for help with Figure 1, and the Servicio de Impuestos Internos (Chilean Tax Authority) for the local Value-added-tax data.

## Funding information

This research was supported by the Agencia Nacional de Investigación y Desarrollo (ANID) Millennium Science Initiative / Millennium Initiative for Collaborative Research on Bacterial Resistance (MICROB-R) [NCN17_081] and Millennium Nucleus for the Study of the Life Course and Vulnerability (MLIV) to EU; Fondo Nacional de Desarrollo Científico y Tecnológico (FONDECYT) [Grant 11191206] to RW; Centro de Desarrollo Urbano Sustentable (CEDEUS), ANID Fondo de Financiamiento de Centros de Investigación en Áreas Prioritarias (FONDAP) [Grant 15110020] to KA; and Centro de Investigación para la Gestión Integrada del Riesgo de Desastres (CIGIDEN), ANID FONDAP [Grant 15110017] to RV.

## Conflicts of interest

The authors declare no conflict of interest.

## Authorship contributions

K.A., E.U., R.V., and R.W. contributed to the design and implementation of the research, to the analysis of the results and to the writing of the manuscript.

## Ethics approval

Our research uses publicly available administrative data collected by the Chilean government; data are aggregated at the municipality-level and subjects cannot be identified directly or through identifiers. It is considered exempt by the Comite Etico Cientifico of Pontificia Universidad Catolica de Chile.

## Supplementary material

**Appendix Table S1.** Regressions results for the effects of mobility and localized lockdown on VAT growth using ordinary least squares regressions and two-stage least squares fixed effects estimates, January to May 2020

| **Panel A: Two stages least squares** | (1) | (2) | (3) |
| --- | --- | --- | --- |
| Dependent variable: VAT growth | OLS 2WFE | OLS 2WFE | IV 2WFE |
| Lockdown as a percentage of a month | -0.131**<br>(0.053) |  |  |
| Log mobility | -0.036<br>(0.140) | 0.043<br>(0.130) | 0.791**<br>(0.032) |
| Observations | 850 | 850 | 850 |
| Adjusted R<sup>2</sup> | 0.351 | 0.349 |  |
| Municipalities | 170 | 170 | 170 |
| **Panel B: First stage** |  |  |  |
| Dependent variable: Log Mobility | (1) | (2) | (3) |
| Lockdown as a percentage of a month |  |  | -0.159***<br>(-5.83) |
| Observations |  |  | 850 |
| Adjusted R<sup>2</sup> |  |  | 0.970 |
| Municipalities |  |  | 170 |

**Notes.** VAT: Value-added tax. Robust standard errors in parenthesis. OLS: Ordinary least squares. 2WFE: two-way fixed effects. Panel A shows the impact of mobility and lockdown on VAT growth using OLS and two-stage least squares fixed effects estimates for the baseline sample. We consider a mobility index as an endogenous variable for January to May that reacts to lockdowns. For each municipality in January and February, we imputed the mean of the daily mobility index during Marc's first fifteen days. Column (1) controls for mobility using the logarithm of the daily average of the mobility index at the municipality level. Column (2) uses the mobility index to explain variations in VAT with OLS 2WFE. In column (3), we use lockdown as an instrumental variable for mobility.

### Mobility

We investigated whether mobility affects economic activity (Table S1). In a simple regression, with and without controlling for lockdowns, the mobility index had a non-significant effect on economic activity. Table S1, column (1) suggests that lockdowns continue having sizable effects even after controlling for mobility. We found no significant effects of mobility on economic activity (Table S1, column 2).

Since lockdowns and mobility could work in the same mechanism, in column (3) of Table S1, we use the method of instrumental variables. This is a method to analyze how lockdown-induced shocks to mobility impact economic activity. For expositional purposes, this is done in two stages. In the bottom panel of column (3), the so-called first-stage has a good fit, meaning that lockdowns impact mobility. On the top panel, the second stage regresses VAT on the lockdown-induced changes in mobility which were calculated in the first stage above. This second stage has a large and significant coefficient of 0.79. To get correct standard errors, these two stages are jointly estimated.

Importantly, this method of instrumental variables tries to decompose the effect on lockdowns on mobility and the subsequent impact of mobility on economic activity. The first coefficient means that a month of lockdown changes monthly mobility by minus 15%. The second coefficient means that lockdown-induced mobility changes VAT by +79%. The multiplication of these effects gives a sense of the net impact of lockdowns on VAT. The multiplication (-0.15 × 0.79) yields a minus 0.11. This is reassuring, since it falls within the range of our baseline estimates in Table 2. Notably, while there could be transmission mechanisms by which lockdowns affect economic activity beyond mobility, these results suggest that mobility is the leading mechanism.

## Notes

### Competing Interest Statement

The authors have declared no competing interest.

### Funding Statement

This research was supported by the Agencia Nacional de Investigacion y Desarrollo (ANID) Millennium Science Initiative / Millennium Initiative for Collaborative Research on Bacterial Resistance (MICROB-R) [NCN17_081] and Millennium Nucleus for the Study of the Life Course and Vulnerability (MLIV) to EU; Fondo Nacional de Desarrollo Cientifico y Tecnologico (FONDECYT) [Grant 11191206] to RW; Centro de Desarrollo Urbano Sustentable (CEDEUS), ANID Fondo de Financiamiento de Centros de Investigacion en Areas Prioritarias (FONDAP) [Grant 15110020] to KA; and Centro de Investigacion para la Gestion Integrada del Riesgo de Desastres (CIGIDEN), ANID FONDAP [Grant 15110017] to RV.

## References

1 Hsiang S, Allen D, Annan-Phan S, Bell K, Bolliger I, Chong T, et al. The effect of large-scale anti-contagion policies on the COVID-19 pandemic. Nature. 2020;584:262–7.

2 Walker PGT, Whittaker C, Watson OJ, Baguelin M, Winskill P, Hamlet A, et al. The impact of COVID-19 and strategies for mitigation and suppression in low- and middle-income countries. Science. 2020;369:413–22.

3 Wiersinga WJ, Rhodes A, Cheng AC, Peacock SJ, Prescott HC. Pathophysiology, Transmission, Diagnosis, and Treatment of Coronavirus Disease 2019 (COVID-19): A Review. JAMA. 2020:Online ahead of print.

4 Flaxman S, Mishra S, Gandy A, Unwin HJT, Mellan TA, Coupland H, et al. Estimating the effects of non-pharmaceutical interventions on COVID-19 in Europe. Nature. 2020;584:257–61.

5 Davies NG, Kucharski AJ, Eggo RM, Gimma A, Edmunds WJ, Jombart T, et al. Effects of non-pharmaceutical interventions on COVID-19 cases, deaths, and demand for hospital services in the UK: a modelling study. The Lancet Public Health. 2020.

6 Lavezzo E, Franchin E, Ciavarella C, Cuomo-Dannenburg G, Barzon L, Del Vecchio C, et al. Suppression of a SARS-CoV-2 outbreak in the Italian municipality of Vo’. Nature. 2020:1-.

7 Hao X, Cheng S, Wu D, Wu T, Lin X, Wang C. Reconstruction of the full transmission dynamics of COVID-19 in Wuhan. Nature. 2020;584:420–4.

8 Cousins S. New Zealand eliminates COVID-19. Lancet. 2020;395:1474.

9 Cowling BJ, Ali ST, Ng TWY, Tsang TK, Li JCM, Fong MW, et al. Impact assessment of non-pharmaceutical interventions against coronavirus disease 2019 and influenza in Hong Kong: an observational study. Lancet Public Health. 2020;5:e279–e88.

10 Pan A, Liu L, Wang C, Guo H, Hao X, Wang Q, et al. Association of Public Health Interventions With the Epidemiology of the COVID-19 Outbreak in Wuhan, China. JAMA. 2020;323:1915–23.

11 Lai S, Ruktanonchai N, Zhou L, Prosper O, Luo W, Floyd J, et al. Effect of non-pharmaceutical interventions to contain COVID-19 in China. Nature. 2020;585:410–3.

12 Rudan I. Answering 20 more questions on COVID-19 (March-April 2020). J Glob Health. 2020;10:020102.

13 Shimizu K, Wharton G, Sakamoto H, Mossialos E. Resurgence of covid-19 in Japan. BMJ. 2020;370:m3221.

14 Ruktanonchai NW, Floyd J, Lai S, Ruktanonchai CW, Sadilek A, Rente-Lourenco P, et al. Assessing the impact of coordinated COVID-19 exit strategies across Europe. Science. 2020.

15 Wise J. Covid-19: Risk of second wave is very real, say researchers. BMJ. 2020;369:m2294.

16 Kissler SM, Tedijanto C, Goldstein E, Grad YH, Lipsitch M. Projecting the transmission dynamics of SARS-CoV-2 through the postpandemic period. Science. 2020;368:860–8.

17 Li Y, Undurraga EA, Zubizarreta JR. Effectiveness of Localized Lockdowns in the SARS-CoV-2 Pandemic. medRxiv. 2020:2020.08.25.20182071.

18 Shea K, Runge MC, Pannell D, Probert WJM, Li S-L, Tildesley M, et al. Harnessing multiple models for outbreak management. Science. 2020;368:577–9.

19 Karatayev VA, Anand M, Bauch CT. Local lockdowns outperform global lockdown on the far side of the COVID-19 epidemic curve. Proc Natl Acad Sci. 2020:202014385.

20 Dong E, Du H, Gardner L. An interactive web-based dashboard to track COVID-19 in real time. Lancet Infect Dis. 2020;20:P533–P4.

21 World Bank. The Economy in the Time of CoVID-19. 2020. Available: https://bit.ly/35B2r3y. Accessed.

22 Baek C, McCrory PB, Messer T, Mui P. Unemployment effects of stay-at-home orders: Evidence from high frequency claims data. Institute for Research on labor and employment working paper. 2020.

23 Lozano-Rojas F, Jiang X, Montenovo L, Simon KI, Weinberg BA, Wing C. Is the cure worse than the problem itself? immediate labor market effects of covid-19 case rates and school closures in the us. National Bureau of Economic Research, 2020 0898–2937.

24 Aum S, Lee SYT, Shin Y. COVID-19 Doesn’t Need Lockdowns to Destroy Jobs: The Effect of Local Outbreaks in Korea. National Bureau of Economic Research, 2020 0898–2937.

25 Gupta S, Montenovo L, Nguyen TD, Rojas FL, Schmutte IM, Simon KI, et al. Effects of social distancing policy on labor market outcomes. National Bureau of Economic Research, 2020 0898–2937.

26 Pfefferbaum B, North CS. Mental Health and the Covid-19 Pandemic. N Engl J Med. 2020.

27 de Girolamo G, Cerveri G, Clerici M, Monzani E, Spinogatti F, Starace F, et al. Mental Health in the Coronavirus Disease 2019 Emergency—The Italian Response. JAMA Psychiatry. 2020;77:974–6.

28 Chowdhury R, Luhar S, Khan N, Choudhury SR, Matin I, Franco OH. Long-term strategies to control COVID-19 in low and middle-income countries: an options overview of community-based, non-pharmacological interventions. Eur J Epidemiol. 2020;epub ahead of print.

29 Mahase E. Covid-19: How does local lockdown work, and is it effective? BMJ. 2020;370:m2679.

30 Bartik AW, Bertrand M, Lin F, Rothstein J, Unrath M. Measuring the labor market at the onset of the COVID-19 crisis. National Bureau of Economic Research, 2020 0898–2937.

31 Forsythe E, Kahn LB, Lange F, Wiczer D. Labor Demand in the time of COVID-19: Evidence from vacancy postings and UI claims. J Public Econ. 2020;189:104238.

32 Goolsbee A, Syverson C. Fear, lockdown, and diversion: Comparing drivers of pandemic economic decline 2020. National Bureau of Economic Research, 2020 0898–2937.

33 Coibion O, Gorodnichenko Y, Weber M. The cost of the covid-19 crisis: Lockdowns, macroeconomic expectations, and consumer spending. National Bureau of Economic Research, 2020 0898–2937.

34 Görg H, Hornok C, Montagna C, Onwordi G. Employment to output elasticities & reforms towards flexicurity: Evidence from OECD countries. 2018.

35 Taylor L. How South America became the new centre of the coronavirus pandemic. New Sci. 2020 10 June.

36 Burki T. COVID-19 in Latin America. Lancet Infect Dis. 2020;20:547–8.

37 de Souza WM, Buss LF, Candido DdS, Carrera J-P, Li S, Zarebski AE, et al. Epidemiological and clinical characteristics of the COVID-19 epidemic in Brazil. Nat Hum Behav. 2020.

38 Moreira RM, Montoya ACV, Araujo SLS, Trindade RA, da Cunha Oliveira D, de Oliveira Marinho G. How prepared is Brazil to tackle the COVID-19 disease? J Glob Health. 2020;10:020321.

39 Taylor L. How Latin America is fighting covid-19, for better and worse. BMJ. 2020;370:m3319.

40 Ministerio de Salud (MINSAL). Plan de acción por coronavirus. 2020. Available: https://www.gob.cl/coronavirus/plandeaccion/. Accessed.

41 Cuadrado C, Monsalves MJ, Gajardo J, Bertoglia MP, Najera M, Alfaro T, et al. Impact of small-area lockdowns for the control of the COVID-19 pandemic. medRxiv. 2020.

42 Bennett M. All Things Equal? Heterogeneity in Policy Effectiveness against COVID-19 Spread in Chile. World Dev. 2020;in press.

43 Gil M, Undurraga EA. CoVID-19 has exposed how ‘the other half’ (still) lives Bull Lat Am Res. 2020;in press.

44 Aguirre P, Asahi K, Díaz-Rioseco D, Riveros I, Valdés R. Medium-run Local Economic Effects of a Major Earthquake. Unpublished Manuscript, Pontificia Universidad Católica de Chile. 2020.

45 Instituto Nacional de Estadísticas (INE). Estimaciones y proyecciones de la población de Chile 1992-2050. 2017. Available: https://www.censo2017.cl/. Accessed.

46 Ministerio de Salud (MINSAL). Cifras Oficiales COVID-19. 2020. Available: https://www.gob.cl/coronavirus/cifrasoficiales/. Accessed.

47 Ministerio de Ciencia T, Conocimiento, e Innovación (Minciencias). Base de datos CoVID-19. 2020. Available: http://www.minciencia.gob.cl/covid19.Accessed.

48 Bavel JJV, Baicker K, Boggio PS, Capraro V, Cichocka A, Cikara M, et al. Using social and behavioural science to support COVID-19 pandemic response. Nat Hum Behav. 2020;4:460–71.

49 Banco Central de Chile. Cuentas Nacionales 2003-2009, Cuadro 1.3. 2010. Available: https://si3.bcentral.cl/estadisticas/Principal1/Informes/CCNN/ANUALES/ccnn_2003_2009.pdf. Accessed.

50 Brzezinski A, Kecht V, Van Dijcke D. The Cost of Staying Open: Voluntary Social Distancing and Lockdowns in the US. Brzezinski, Adam, Kecht, Valentin, and Van Dijcke, David (2020)” The Cost of Staying Open: Voluntary Social Distancing and Lockdowns in the US” Economics Series Working Papers. 2020;910.

51 Frenier C, Nikpay SS, Golberstein E. COVID-19 Has Increased Medicaid Enrollment, But Short-Term Enrollment Changes Are Unrelated To Job Losses: Study examines influence COVID-19 may have had on Medicaid enrollment covering the period of March 1 through June 1, 2020 for 26 states. Health Aff. 2020:10.1377/hlthaff. 2020.00900.

52 Case A, Deaton A. Deaths of Despair and the Future of Capitalism: Princeton University Press; 2020.

53 Levy-Yeyati E, Valdés R. COVID-19 in Latin America: How is it different than in advanced economies? In: Djankov S, Panizza U, editors. COVID-19 in Developing Economies CEPR Press VoxEU.org eBook; 2020.

